# Development of Loop-mediated Isothermal Amplification (LAMP) Assays Using Five Primers Reduces the False-positive Rate in COVID-19 Diagnosis

**DOI:** 10.1101/2022.10.18.22281181

**Authors:** Galyah Alhamid, Huseyin Tombuloglu, Ebtesam Al-Suhaimi

## Abstract

The reverse-transcription loop-mediated isothermal amplification (RT-LAMP) is a cheaper and faster testing alternative for detecting SARS-CoV-2. However, high false-positive rate due to misamplification is one of the major limitations. To overcome misamplifications, we developed colorimetric and fluorometric RT-LAMP assays. The assay performances was verified by the gold-standard RT-qPCR technique on 150 clinical samples. Compared to other primer sets with six primers (*N, S*, and *RdRp*), E-ID1 primer set, including five primers, performed superbly on both colorimetric and fluorometric assays, yielding sensitivities of 89.5% and 100%, respectively, with a limit of detection of 20 copies/µL. The colorimetric RT-LAMP had a specificity of 97.2% and an accuracy of 94.5%, while the fluorometric RT-LAMP obtained 96.9% and 98%, respectively. No misamplification was evident even after 120 minutes, which is crucial for the success of this technique. These findings are important to support the use of RT-LAMP in the healthcare systems in fighting COVID-19.

## Introduction

The coronavirus disease (COVID-19), caused by the severe acute respiratory syndrome coronavirus 2 (SARS-CoV-2), continues to cause rapid infections and deaths since its emergence over two years ago, reaching up to 604 million cases and over 6.4 million deaths worldwide as of August 2022, according to world health organization (WHO) (*COVID Live -Coronavirus Statistics -Worldometer*, 2022). Efforts to control the spread of this disease are continuous. However, the emergence of new variants with high mutation rates that cause increased infectivity and decreased effectiveness of vaccines, such as Omicron (B.1.1.529), hinders the efficient control of this pandemic. Reverse-transcription quantitative polymerase chain reaction (RT-qPCR) is the accepted gold standard technique to diagnose this disease. However, it requires sophisticated equipment, a laboratory environment, and expert personnel to perform the tests. These requirements prevent rapid and expanded testing, especially in regions with lower resources. Therefore, researchers are investigating alternative and affordable point-of-care (POC) testing methods that can diagnose COVID-19 with high sensitivity. One of the molecular techniques that meet these criteria is reverse-transcription loop-mediated isothermal amplification (RT-LAMP), which uses four to six primers to target and amplify a specific gene region. The amplification reaction in this technique takes place isothermally in a single tube without requiring bulky instruments and provides simple detection techniques, being also cheaper and faster than the gold standard (Notomi, 2000, Notomi et al., 2015; (Alhamid and Tombuloglu, 2022). The efficiency of RT-LAMP in COVID-19 diagnosis was demonstrated by many researchers (Aoki et al., 2021, Bokelmann et al., 2021, Dao Thi, 2020, de Oliveira Coelho et al., 2021, Lu et al., 2020; Alhamid et al., 2022), and many validated the performance of FDA-approved colorimetric RT-LAMP kits for emergency use authorization (Baba et al., 2021, Promlek et al., 2022, *Color SARS-CoV-2 RT-LAMP Diagnostic Assay -EUA Summary*, 2021). However, because of the number of primers, the most common limitation of the RT-LAMP technique is the misamplifications that arise from unwanted secondary structures. Even with the careful primer design and the availability of programs that check for dimer and hairpin structures, there is no guarantee that these structures will not be formed practically. In particular, current protocols recommend that the reaction time not exceed 30 minutes to avoid misleading results (Amaral et al., 2021, Aoki et al., 2021, de Oliveira Coelho et al., 2021). This makes it challenging to detect samples with low copy numbers and reduces the efficiency of the RT-LAMP technique.

To address this issue, we hypothesize that a lower number of LAMP primers will reduce the false positivity rate, which is the most common limitation of the LAMP technique (Meagher et al., 2018, Odiwuor et al., 2022). The slower amplification rate can be improved by optimizing the performance of the developed colorimetric and fluorometric assays via increasing the enzyme’s concentration or adding primer binding enhancers like guanidine hydrochloride, as shown elsewhere (Dudley et al., 2020, Lu et al., 2022, Zhang, 2020b). In this work, we demonstrate the efficiency of using five primers to reduce misamplifications by comparing them with five primer sets targeting different genes such as *RdRP, S*, and *N*. The optimized protocol using five primers (E-ID1) eliminates the misamplifications, thus improving the detection’s sensitivity and efficacy. By the time of writing this manuscript, there were no studies that discussed the efficiency of primer sets with five primers in the colorimetric and fluorometric detection of SARS-CoV-2, which will be comprehensively addressed here.

## Materials and methods

### Alignment of SARS-CoV-2 genome sequences and RT-LAMP primer design

Genomic sequences of all SARS-CoV-2 types that have been sequenced worldwide were downloaded from the database of GISAID (Global Initiative on Sharing All Influenza Data, <https://www.gisaid.org>) and NCBI GenBank <https://www.ncbi.nlm.nih.gov/genbank/>. At the time of writing, 12,848,795 hCoV-19 genomes were submitted worldwide. The downloaded genome sequence information was sampled from different continents and included the globally dominant variants such as alpha (B.1.1.7, Q.1-Q.8), beta (B.1.351, B.1.351.2, B.1.351.3), gamma (P.1, P.1.1, P.1.2), delta (B.1.617.2), and Omicron (B.1.1.529) to identify the most conserved regions for RT-LAMP primer design. This way, the designed assay would target the viral gene as sensitive, specific, and accurate as possible, regardless of the variant. Also, whole genome sequences of other SARS-CoV like AY278491.2, AY502924.1, AY502927.1, AY559094.1, AY613947.1, and NC_004718.3 were downloaded from the NCBI database. Then, a comparative analysis was made by aligning the multiple sequences at the base level with the bioinformatics program Clustal Omega <https://www.ebi.ac.uk/Tools/msa/clustalo/>. Afterward, the viral target regions were selected in conformity with COVID-19 testing directives of the CDC, WHO, and EU Commission. The mutation sites were identified using JalView (v2.11.1.3) program (Waterhouse et al., 2009). The Conserved regions specific for SARS-CoV-2 but not for the other SARS-COV species were selected for primers’ binding sites. All RT-LAMP primer sets, each containing five to six primers, were designed using PrimerExplorer V5 program <https://primerexplorer.jp/e> that target the conserved region in the *N, S, RdRp*, and *E* genes. In addition, the OligoAnalyzer tool from Integrated DNA Technologies (IDT) <https://eu.idtdna.com/pages/tools/oligoanalyzer> was used to check that each primer set would not form unwanted secondary structures like homodimer, hetero-dimer, and hairpins. The selected primers were synthesized by Alpha DNA (Montreal, Canada) <http://www.alphadna.com>. These primers were lyophilized and desalted. All the designed primer sets were tested on SARS-CoV-2 positive control (PC)—a mixture of verified high SARS-CoV-2-loaded specimens—and non-template control (NTC)—distilled water—and the ones that showed the best performance were chosen for further testing and analysis. In 25 µL reaction volume, 10x primers’ concentrations in a primer mix were as follows: 2 µM for F3 and B3, 4 µM for LF and LB, and 16 µM for FIP and BIP. Sequences of all primer sets are shown in Table 1.

**Table 1.**
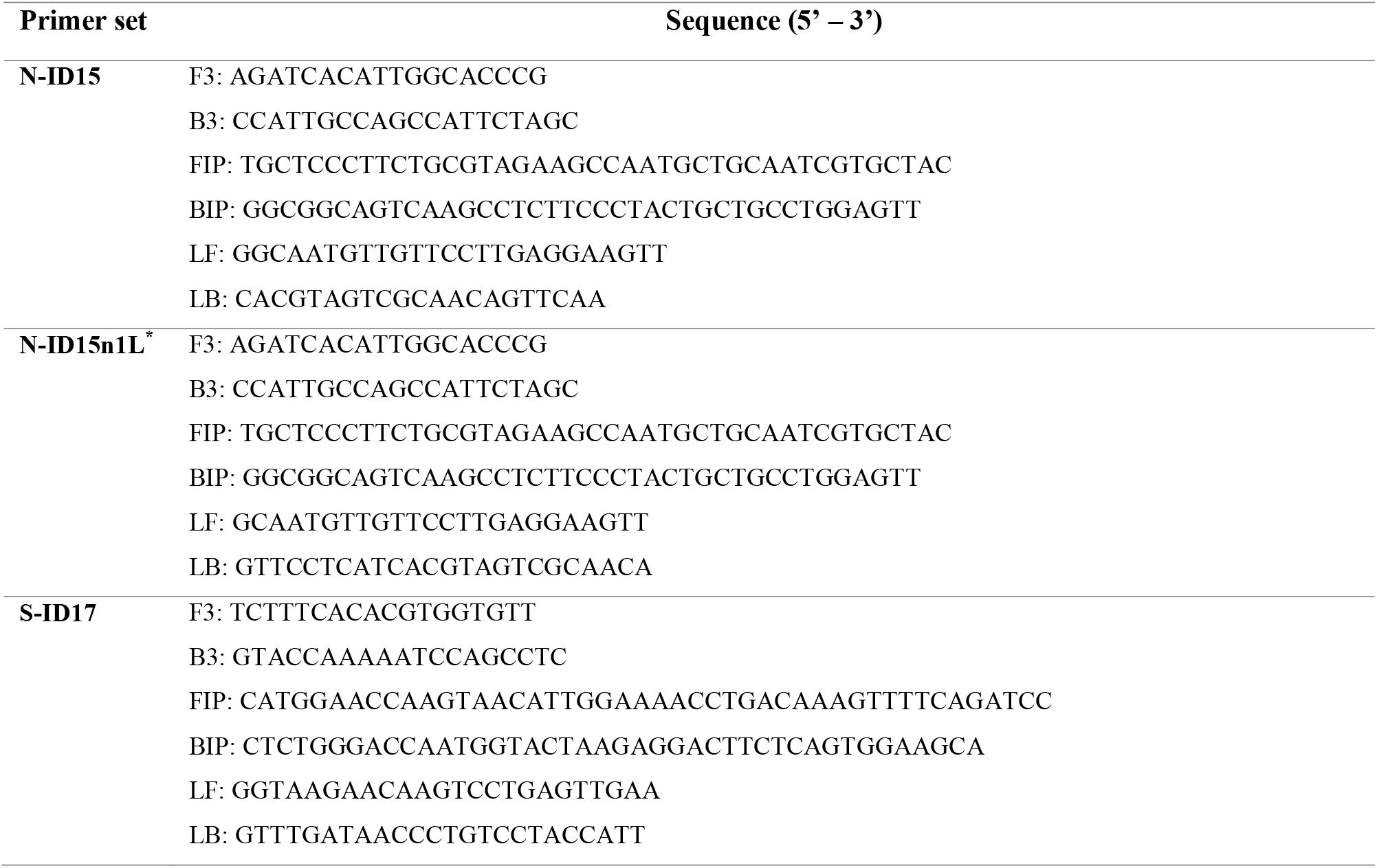
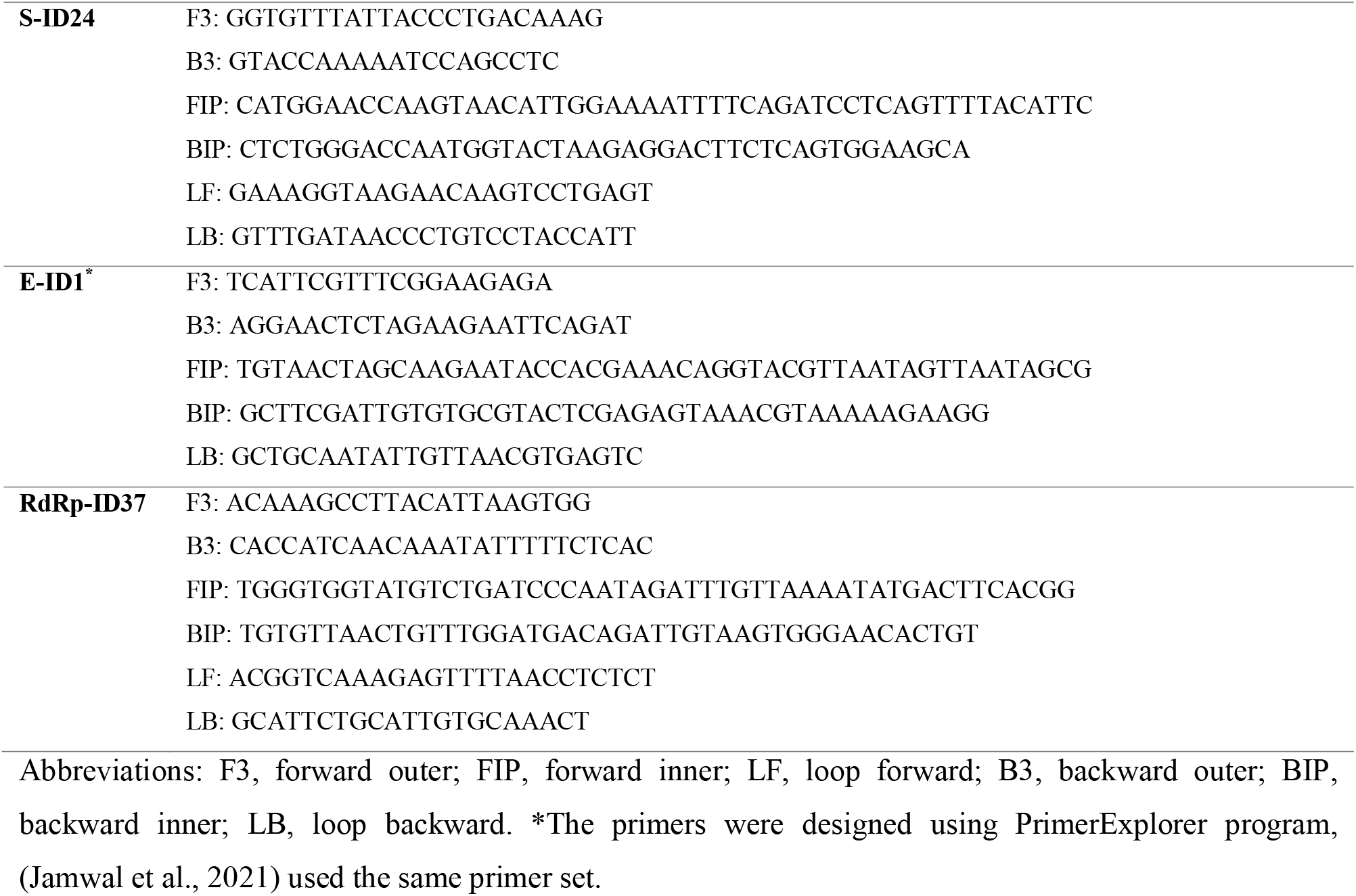
Sequences of each RT-LAMP primer set used in this study.

### Fluorometric RT-LAMP reaction

The Fluorometric RT-LAMP reactions were carried out in 25 μL reaction volume. Two brands were tested in the fluorometric observation to choose the most compatible reagents that show the best performance: WarmStart^®^ (New England BioLabs) and LavaLAMP™ (Lucigen). Unless otherwise stated, a LavaLAMP™ master mix contained 2.5 µL 10x LavaLAMP™ RNA buffer, 1.25 µL 100 mM MgSO_4_, 2 µL 10 mM dNTP solution, 1 µL 20x Green Fluorescent Dye, 1 µL LavaLAMP™ RNA enzyme (Lucigen), 2.5 µL primer mix, 5 µL template (or dH_2_O for NTC), and dH_2_O up to 25 µL. In the fluorometric assay using WarmStart^®^ reagents, the master mix included 2.5 µL 10x isothermal amplification buffer, 1.5 µL 100 mM MgSO_4_, 3.5 µL 10 mM deoxynucleotide (dNTP) solution, 0.5 µL 50x LAMP fluorescent dye, 8000 U/mL Bst 2.0 WarmStart^®^ DNA polymerase, 0.5 µL WarmStart^®^ RTx reverse transcriptase (New England BioLabs), 2.5 µL primer mix, 2 µL template (or dH_2_O for NTC), and dH_2_O up to 25 µL. Reactions were performed in a 7500 Fast Real-Time PCR System, Thermo Fisher Scientific (Waltham, MA, USA) <https://www.thermofisher.com>, at 65 °C for Warmstart^®^ and 70 °C for LavaLAMP™ for 80 cycles, 45-60 seconds using FAM filter as the reporter dye channel.

### Colorimetric RT-LAMP detection

For a 25 µL total reaction volume, unless otherwise specified, the colorimetric RT-LAMP mixture is composed of 12.5 µL WarmStart^®^ Colorimetric LAMP 2x master mix with UDG (New England BioLabs), 2.5 µL 10x primer mix, 2 µL template (or dH_2_O for NTC), and dH_2_O up to 25 µL. The mixture is incubated in a water bath, Thermo Fisher Scientific (Waltham, MA, USA) <https://www.thermofisher.com>, set at 65 °C for 60 minutes or until a distinguishable color change is observed.

### Agarose gel electrophoresis

Positive or negative amplifications upon RT-LAMP reactions were validated by loading RT-LAMP products in 2% agarose gel prepared with VisualaNA (A) DNA Stain from Molequle-On (Auckland, New Zealand) <http://molequle-on.com> and run in electrophoresis unit (Analytik Jena) for 45 minutes operating at 100 V. Then, the gel is visualized under a UV-trans illuminator (ChemiDoc™ XRS+ System with Image Lab™ Software, Bio-Rad, USA). Successful amplifications in positive samples were visualized as ladder-type DNA bands.

### Collection of samples and validation of results

Multiple validations were conducted on real clinical specimens in collaboration with the Microbiology Department of King Fahd University Hospital (KFUH), Dammam, which is authorized to store and analyze SARS-CoV-2 samples from patients. A sufficient number of samples (150 validated SARS-CoV-2 positive or negative RNA samples) were simultaneously tested and validated with the herein developed colorimetric RT-LAMP assay, fluorometric RT-LAMP assay, and other commercially or in-house developed RT-qPCR kits (Tombuloglu et al., 2021, Tombuloglu et al., 2022). The limit of detection (LoD) was determined using serial dilutions of synthetic SARS-CoV-2 RNA control (Twist synthetic RNA control 51 (EPI_ISL_7718520), Twist Bioscience, USA). The specificity was determined by testing other respiratory viruses’ RNA extracted from clinical samples, including parainfluenza virus 3, enterovirus, rhinovirus, human metapneumovirus A+B, parainfluenza virus 4, bocavirus, and coronavirus 229 E. Eventually, the following results for further validation were obtained: 1) RT-qPCR results from the abovementioned hospitals, 2) RT-qPCR results from *in situ* lab, 3) fluorescent RT-LAMP, 4) the colorimetric RT-LAMP, and 5) agarose gel electrophoresis.

### RT-qPCR assay

Since RT-qPCR assay is accepted as the gold standard method, the positivity or negativity of the collected specimens was tested to verify the RT-LAMP results. RNA samples were used as template by targeting at least two viral genes (*RdRp, N*, and *E*) and a human *RP* gene as the internal control, as described earlier (Tombuloglu et al., 2021, Tombuloglu et al., 2022). In addition, no-template control (NTC) was used to detect any possible misamplifications. The reactions were run in a real-time PCR instrument (Applied Biosystems™ real-time PCR 7500).

The samples having a threshold cycle (Ct or Cq) score over 37 (Ct > 37) were considered negative. The SARS-CoV-2 positive specimens were identified if the Ct number was ≤ 37 with a sigmoidal amplification curve.

### Ethical approval

The study is approved by the Institutional Review Board (IRB) at Imam Abdulrahman bin Faisal University (IAU) with an IRB number of IRB-2020-13-406. All methods were carried out in accordance with relevant guidelines and regulations. The de-identified samples left over after completion of diagnostic tests were used; hence this study requires no consenting as per institutional ethics committee regulations and informed consent.

### Statistical analysis

For the colorimetric quantification, the difference between the positive and negative samples’ color development was determined spectrophotometrically by the mean values of the optical density (ΔOD). This was done by transferring equal amounts of five positive and five negative clinical samples in a 96-well cell culture plate (Thermofisher Scientific) and reading absorbance values at wavelengths 434 and 560 nm using a plate reader (BioTek Synergy HTX microplate reader, Agilent). The difference in optical densities was considered statistically significant if the *p*-value <0.05 in an unpaired *t*-test performed using GraphPad Prism 9.0 (GraphPad Software, USA).

## Results

### RT-LAMP primers performance

The performance of the five primer sets targeting *N, E, S*, or *RdRp* genes was tested on positive (PC) and non-template controls (NTC). In the colorimetric identification, the color in all tubes was observed at 0 minutes (before reaction) and then subsequently every ten minutes. According to the results shown in Figure 1, primer sets N-ID15n1L and RdRp-ID37 showed a red-to-yellow color difference in positive reactions at 40 minutes. Primer set E-ID1 started to develop a color change at 50 min (without optimization), while most primer sets started to show misamplifications in NTC at this time. The reaction was terminated after 60 minutes due to developing false positivity in NTC in all the sets except E-ID1.

**Figure 1.**
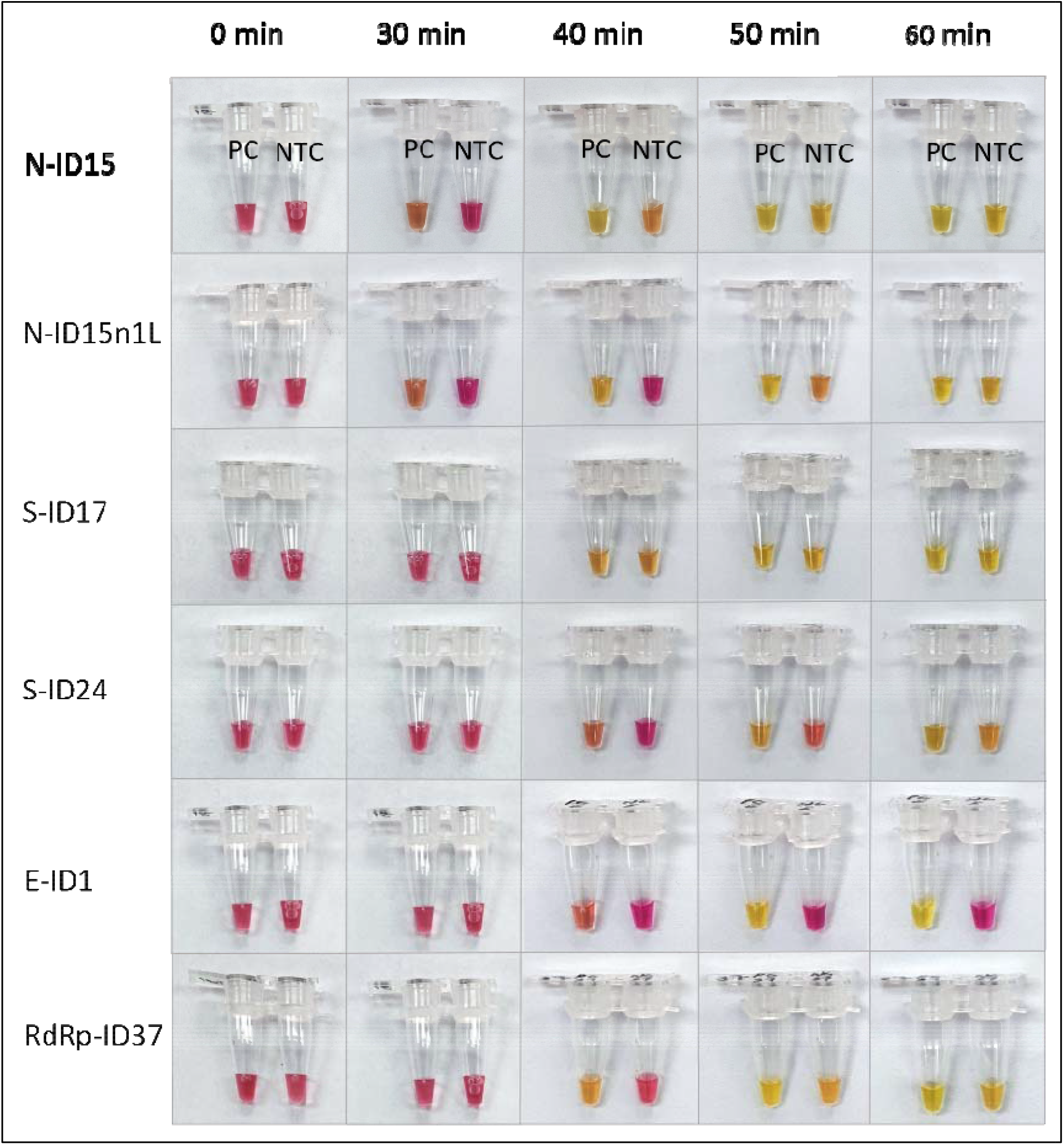
Colorimetric test using all primer sets on positive controls (PC) and non-template controls (NTC). A visible color change from red to yellow develops in PC tubes in th colorimetric identification, and negatives remain red.

Depending on the primer set, the appearance time of false-positive results differs. Since misamplification is evident in the NTC tubes of N-ID15 and S-ID17 for around 40 min, it is not recommended to incubate the reaction for longer than 40 minutes. However, extending the time over 120 minutes did not lead to misamplifications for the E-ID1 set harboring five primers. Therefore, the E-ID1 primer set was selected for further testing.

### Optimization of the colorimetric and fluorometric RT-LAMP assays

Colorimetric and fluorometric RT-LAMP assays were developed and optimized to enhance the performance of the *E* gene primers. The optimization includes addition of guanidine hydrochloride, testing different DNA polymerases (Bst 2.0 *vs* Bst 3.0 versions), and adjusting the optimum reaction temperature. In contrast to the detection time before optimization, the colorimetric assay showed an earlier color change within 30 minutes (Figure 2(a)). Also, this improvement was seen in the fluorometric RT-LAMP assay, where it detected all positive samples in less than 60 minutes, an average of around 40 min, compared to its performance before optimization (Figure 2(b, c)). Supplementary Table S1 compares of the assay’s detection speed and sensitivity on different clinical samples.

**Figure 2.**
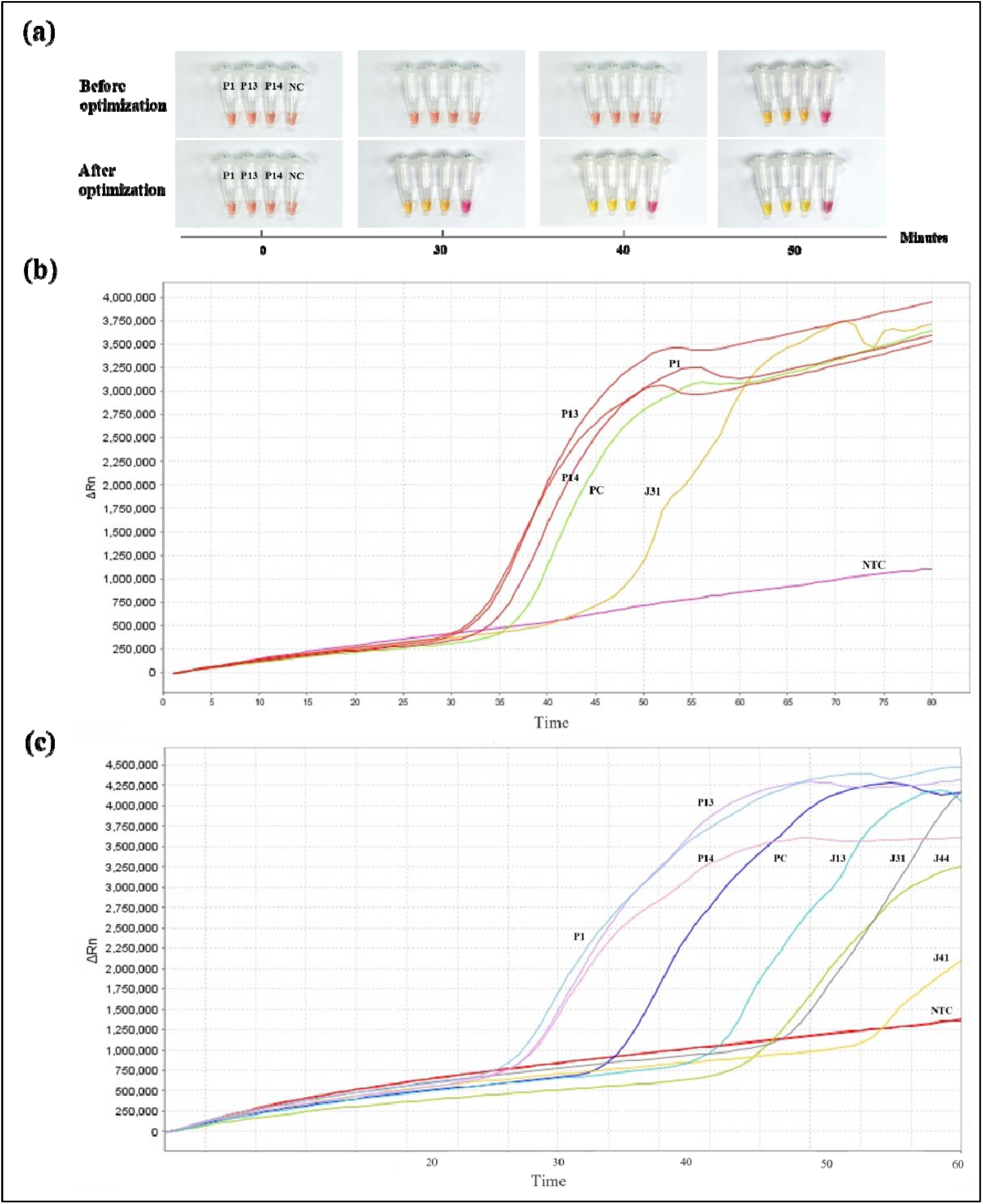
**(a)** Colorimetric RT-LAMP using E-ID1 primer set on clinical samples before and after optimization. This colorimetric assay started to detect the positive samples within 30 minute compared to 50 minutes before optimization. **(b)** Fluorometric RT-LAMP before and ***(c)*** after optimized using LavaLAMP™ reagents on different clinical samples, where the detection enhancement can be evident.

Bst 3.0 enzyme demonstrates improvements in amplification performance and high reverse transcriptase activity, allowing single enzyme RT-LAMP reactions. By comparing with Bst 2.0 DNA polymerase in the fluorometric RT-LAMP, the enzyme that shows the best performance with the E-ID1 primer set was chosen for further analysis. Both enzymes were added at the same concentration on PC and NTC triplicates. However, positive amplifications appeared around 17 minutes using Bst 2.0 enzyme, while with Bst 3.0, PC amplified after 21 minutes (Figure S1).

The effect of guanidine hydrochloride (GuHCl) on the performance of E-ID1 primer set in both colorimetric and fluorometric RT-LAMP reactions was tested since it was reported to enhance the detection speed and efficiency (Dudley et al., 2020, Lu et al., 2022, Zhang, 2020b). Figure 3 (a) shows two sets of colorimetric RT-LAMP with and without the addition of GuHCl (40 mM) on positive clinical specimens, in addition to PC and NTC. The colors started to develop in the positive samples after 24-27 minutes. In reactions with GuHCl, the sample J49 started to show a color change after 27 minutes, and the light-yellow color fully developed after 30 minutes. On the other hand, this sample’s color started to change after 35 minutes without adding GuHCl, which fully developed after 40 minutes (Figure 3 (b)). The effect of GuHCl was also tested fluorometrically on PC and NTC triplicates, and it indeed improved the detection speed. No misamplification in either test was evident upon 120 min.

**Figure 3.**
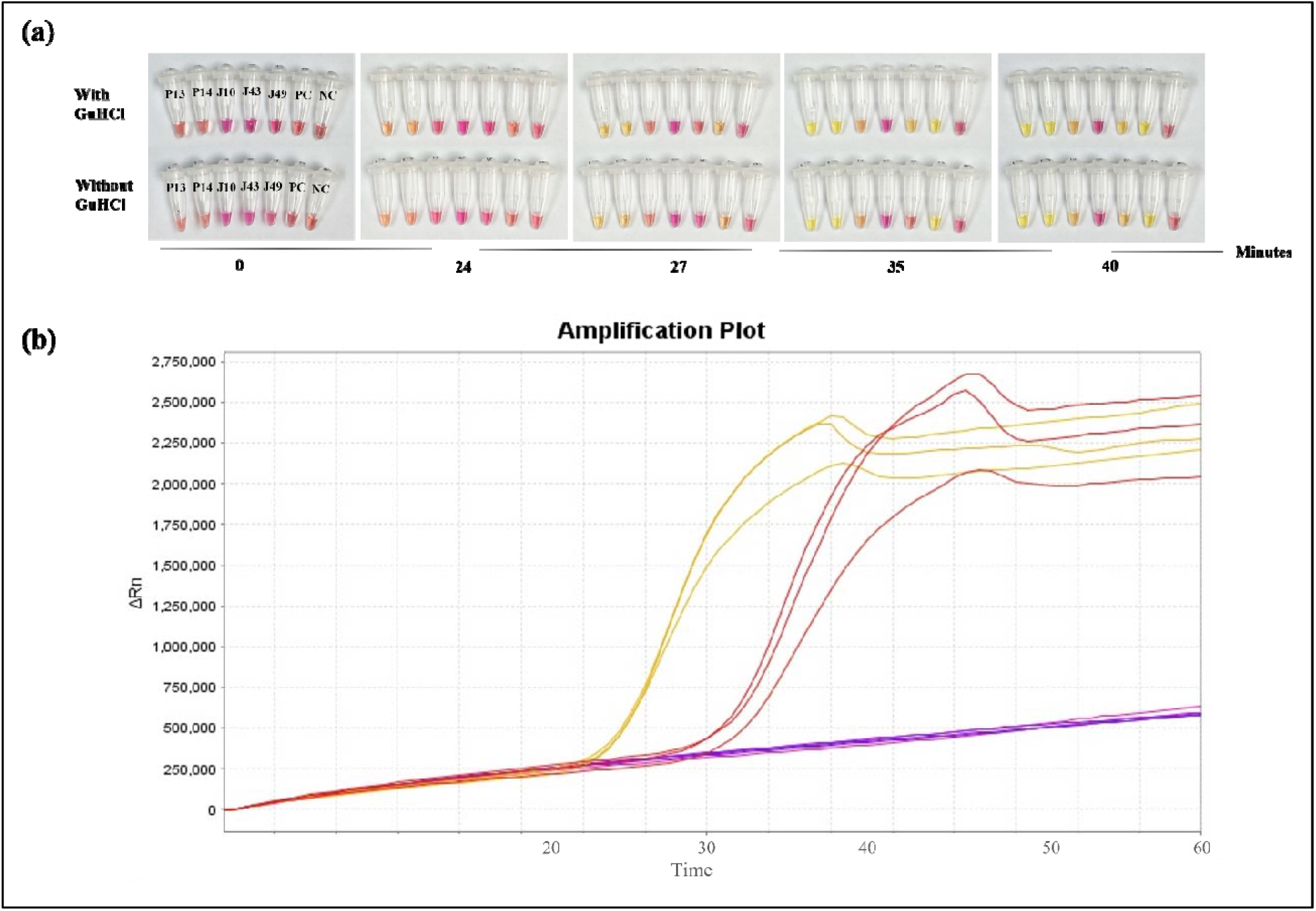
Testing the effect of adding 40 mM guanidine hydrochloride (GuHCl) in **(a)** colorimetric RT-LAMP reaction on positive clinical samples (P13, P14, J10, J49) and an inconclusive sample (J43). This addition improved the detection speed in one sample, where it started to change color eight minutes earlier compared to reactions without GuHCl. The inconclusive sample J43, however, remained negative in both. **(b)** Fluorometric RT-LAMP PC and NTC triplicate test with (yellow) and without GuHCl (red). All PC triplicates with GuHCl amplified earlier than those without. There were no amplifications in NTC in both colorimetric and fluorometric tests, suggesting that GuHCl indeed increases the detection speed without causing misamplification in NTC.

### Limit of detection of *E* gene primers

SARS-CoV-2 synthetic RNA control (Twist synthetic RNA control 51 (EPI_ISL_7718520), Twist Bioscience, USA) was serially diluted (1, 10^1^, 10^2^, 10^3^, 10^4^, 10^5^, and 10^6^ times) to determine the limit of detection (LoD) of RT-LAMP reactions. Figure 4 (a) shows the LoD of the colorimetric RT-LAMP assay targeting the *E* gene. The color development to yellow is prominent in 1, 10^1^, 10^2^, and 10^3^-times dilutions, corresponding to 500 copies/reaction volume (25 µL) or 20 copies/ µL for the 10^3^-times dilution. After the reaction, the reaction mixture of each dilution was loaded in 2% agarose and visualized under a UV trans-illuminator to further check if band-pattern intensities decreased with reduced viral loads (Figure 4 (b)). The results indicate a ladder-type banding pattern in the detected dilutions, while no bands appeared in the others. Agarose gel electrophoresis and the colorimetric identification are in line.

**Figure 4.**
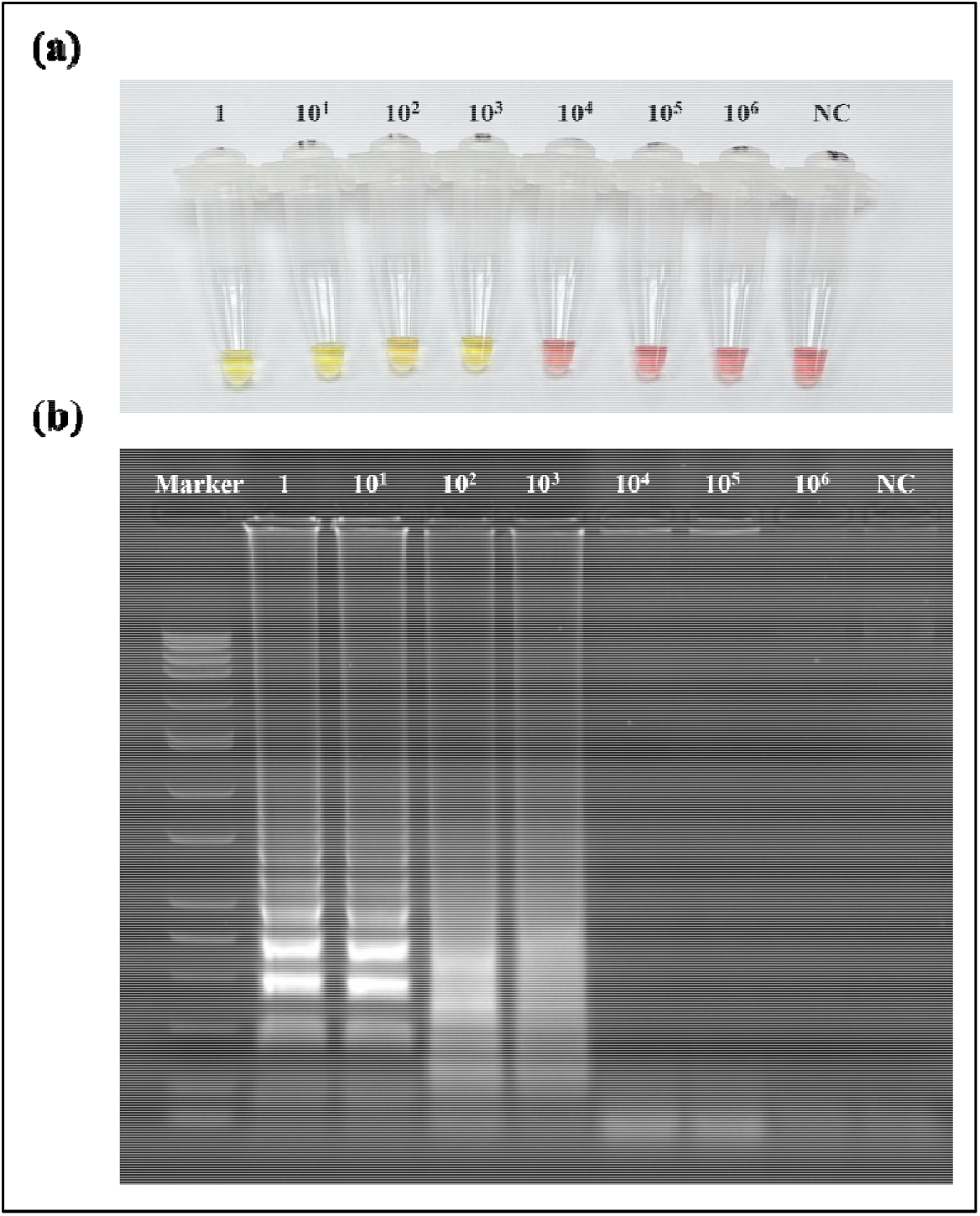
The limit of detection (LoD) of *E* gene primers tested in the colorimetric RT-LAMP assay on serially diluted SARS-CoV-2 synthetic RNA control **(a)**. The assay detected up to 10^3^ dilutions, which correspond to 500 copies/reaction volume (25 µL) or 20 *copies/µL*. **(b)** Validation of the colorimetric LoD results via loading the RT-LAMP product post-reaction in 2% agarose gel showing band patterns of the detected diluted samples.

### RT-LAMP assays on clinical samples

The performance of the E-ID1 primer set was colorimetrically and fluorometrically tested on clinical specimens, as depicted in Figure 5 (a). These results were then validated using agarose gel electrophoresis (Figure 5 (c)). In the colorimetric RT-LAMP reaction, the tubes were incubated in a thermal block set at 65 °C, and the positive clinical specimens started to show a color change after 20 minutes, while the color difference between positives and negatives was the clearest after 30 minutes. The reaction was then terminated after 40 minutes. Eventually, there was 94.5% agreement between the colorimetric RT-LAMP and RT-qPCR results.

**Figure 5.**
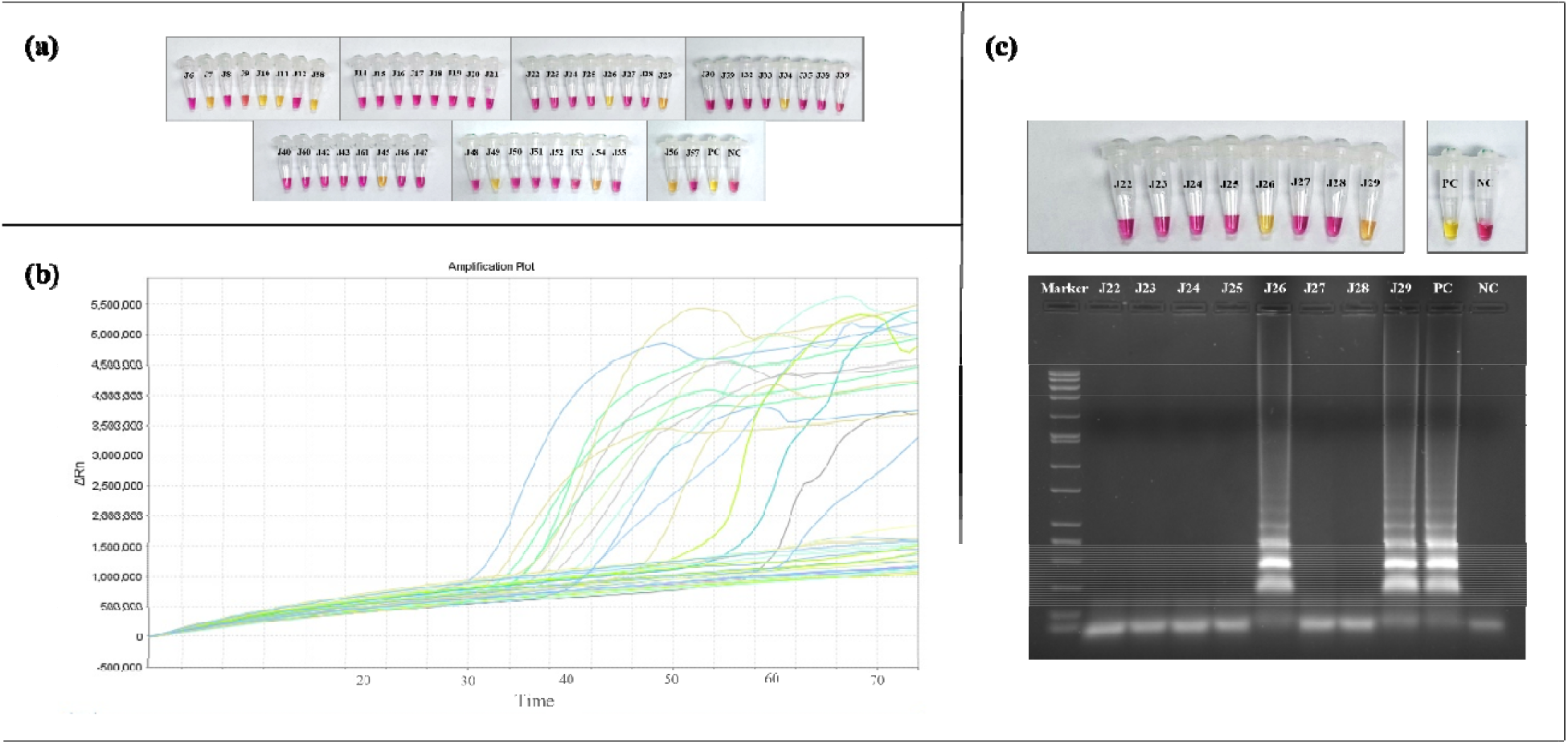
Optimized **(a)** colorimetric and **(b)** fluorometric RT-LAMP assays tested on SARS-CoV-2 clinical samples show agreement in the detection sensitivity. **(c)** Loading eight samples in 2% agarose stained with a DNA stain, a 0.05-10 Kb DNA ladder was used as a marker, where positive samples show DNA bands under UV transilluminator.

Similarly, clinical samples were fluorometrically tested using the optimized RT-LAMP protocol (Figure 5(b)). The reaction was placed at 70 °C in a thermal cycler for 75 minutes. This fluorometric assay had 98% agreement with RT-qPCR results.

### Sensitivity and specificity of RT-LAMP Assays

A total of 150 clinical specimens were tested in both assays. Accordingly, the colorimetric RT-LAMP assay had a sensitivity of 89.5%, specificity of 97.2%, and accuracy of 94.5%. The positive percent agreement (PPA) with the RT-qPCR was calculated to be 94.4%, while the negative percent agreement (NPA) was 94.6%. In the fluorometric detection, the results revealed 100% sensitivity, 96.9% specificity, and 98% accuracy. The fluorometric RT-LAMP assay had a PPA of 94.7% and an NPA of 100%.

The color development in the positive and negative samples was quantified post-reaction using the spectrophotometer by measuring the absorbance at 434 and 560 nm wavelengths. Figure 6 (a) shows that the difference in optical densities (ΔOD) between the positive yellow and negative red samples is statistically significant (p<0.0001, 95% CI). Among the tested samples, the samples having high viral loads were detected in ∼20 minutes, while those with low viral loads took up to 75 minutes (Figure 6 (b, c)). On average, the detection time was calculated as 37 minutes. Besides, 90% of SARS-CoV-2 positive samples were detected within 50 minutes (Figure 6 (c)).

**Figure 6.**
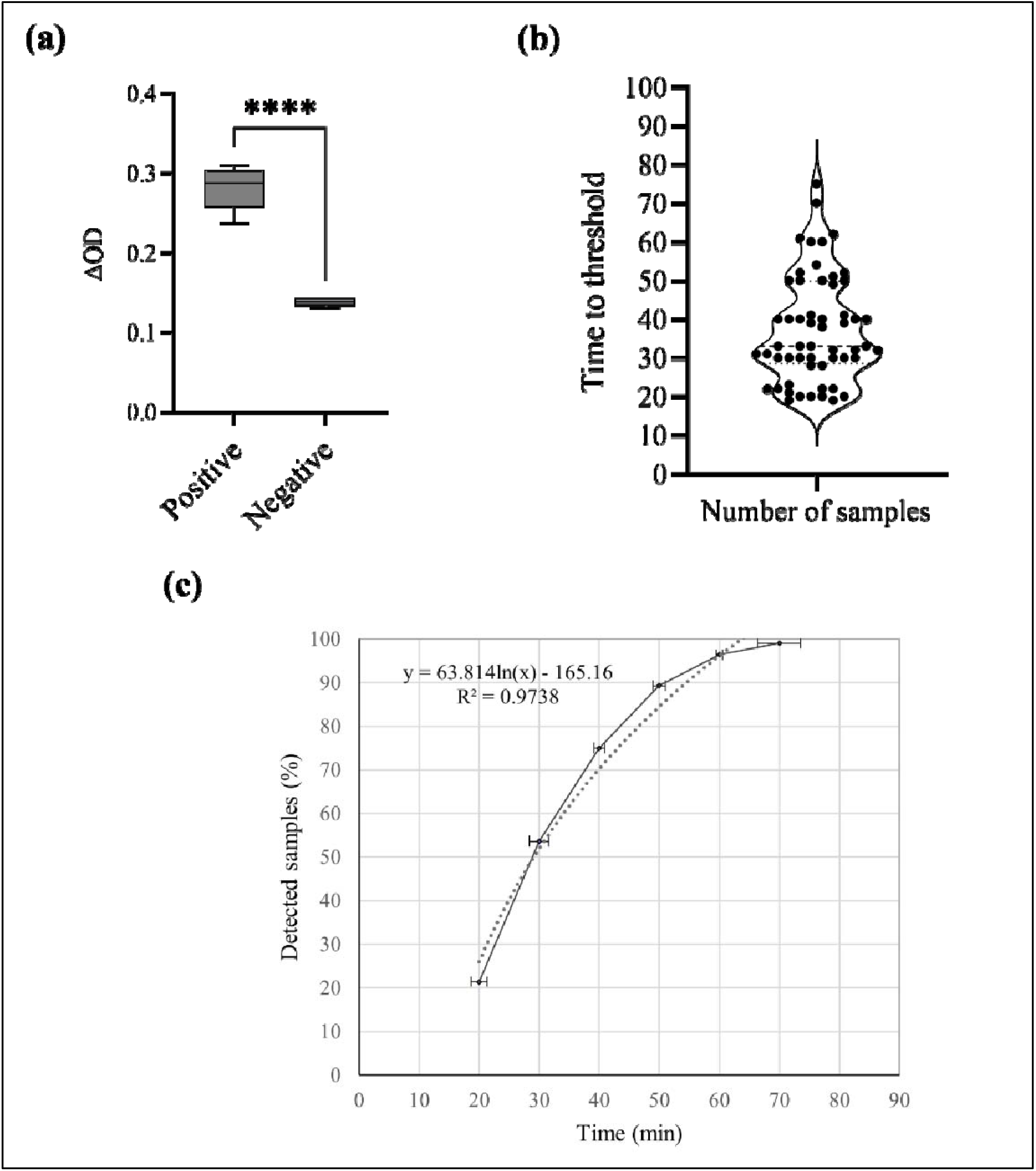
(a) Quantification of the color change between positive and negative samples by the spectrophotometric measurement of optical density (ΔOD) at 434 and 560 nm. The line inside the box represents the median and the whiskers extend to the maximum and minimum values. The four asterisks (*) correspond to p<0.0001, which is a statistically significant difference (unpaired t-test, p<0.05, 95% CI). **(b)** Time to detection threshold of all positive samples. The dash line indicates the mean (37 minutes) and the dotted line represents the median (33 minutes). **(c)** Percentage of total positive samples detected plotted against time.

In addition, the inclusivity of the E-ID1 primer set was tested *in-silico* by aligning the five VOC with other SARS viruses. These primers bind to the conserved regions in all SARS-CoV-2 variants, including the recent Omicron but are uncommon for the other SARS (Figure S3). In addition, the cross-reactivity of E-ID1 primers was tested *in-vitro* against the most common respiratory viruses, including parainfluenza virus 3, enterovirus, rhinovirus, human metapneumovirus A+B, parainfluenza virus 4, bocavirus, and coronavirus 229 E. The colorimetric assay only detected SARS-CoV-2 but none of the other respiratory viruses, which indicates high specificity against the diagnosis of COVID-19 infection (Figure S4).

## Discussion

Diagnostic tests are the first line of defense against the uncontrolled spread of pandemics. Such tests must be accessible, fast, and simple with high diagnostic accuracy to be used for POC in resource-limited areas. Aside from the gold standard RT-qPCR, alternative molecular and serological techniques are being widely investigated to develop fast, accurate, and cost-effective tests to efficiently diagnose pathogenic diseases, including the recent COVID-19. The colorimetric RT-LAMP technique’s simplicity allows for its use as a POC testing alternative for large-scale screening. In this work, two RT-LAMP assays were developed to target and detect the *E* gene of SARS-CoV-2. The colorimetric RT-LAMP holds the advantage of simple and cost-effective reaction conditions, in addition to the visual interpretation of the results. Its diagnostic accuracy, however, must be comparable to the gold standard RT-qPCR to consider this technique reliable to avoid false results and their undesired consequences. In the colorimetric detection, 94.5% of the results were found to be consistent with RT-qPCR, and three samples contradicted it, hence were considered inconclusive. We also monitored the rate of color change in positive specimens throughout incubation times at 0, 20, 30, and 40 minutes to detect the minimum time when the color between positives and negatives is distinguishable. It was observed that the color in high viral load samples started to change after 20 minutes, and lower viral load samples began to change to yellow after 30-40 minutes. Over 50% of positive samples changed to yellow at 30 minutes (Figure 6). This color difference between the positive and negative color was statistically significant (p<0.0001, 95% CI) as the ΔOD of both were measured spectrophotometrically. Overall, the sensitivity of the colorimetric RT-LAMP reaction greatly depends on the assay’s LoD, sample quality, viral load, and time of sample collection from the disease onset (Promlek et al., 2022).

The fluorometric RT-LAMP assay also showed a great performance when testing the clinical samples, where 98% of the results agreed with RT-qPCR. The fluorometric and colorimetric results agree in all tested samples except the inconclusive samples—namely J20, which was fluorometrically negative but colorimetrically positive; and J43, which was fluorometrically positive but colorimetrically negative— (Figure S2). This difference in the results is due to the low viral load in these samples (Ct >30). It can also be caused by insufficient pH reduction to cause a color change to yellow. The solution’s pH is also affected by the elution buffer used during the sample’s RNA extraction process (Aoki et al., 2021). It should be noted that one clinical specimen (J15) was detected in both colorimetric and fluorometric RT-LAMP but was negative in RT-qPCR. Accordingly, both assays had higher sensitivity than RT-qPCR in detecting SARS-CoV-2 in this specimen.

Among the designed primer sets, the *E* gene primers target conserved, non-mutated regions of the most recent VOC but are uncommon for other SARS, based on our *in-silico* analysis. So, this assay is expected to detect any SARS-CoV-2 variant, including the heavily mutated Omicron. Indeed, even with Omicron specific point mutation (C/T) at FIP binding site, this primer set efficiently detected Omicron-infected samples along with the synthetic copy of Omicron (Twist Bioscience). The five primers in this set caused a delayed detection time in positive specimens compared to other primer sets composed of six primers, as demonstrated in Figure 1. However, other primer sets (N-ID15, N-ID15n1L, S-ID17, S-ID24, and RdRp-ID37) had non-specific amplification in NTC appeared in 40 minutes (N-ID15 and S-ID17). The misamplification was clear in all tested primer sets within 50-60 minutes, except the E-ID1 set. Regardless of the late color development, the E-ID1 primer set showed high stability at a longer reaction time when the incubation lasted up to two hours (data not shown). For this reason, we tested tripling the concentration of Bst 2.0 DNA polymerase to compensate for the smaller number of primers. Increasing the enzyme concentration up to three times normally causes non-specific amplification in NTC due to the number of primers and high polymerase activity; however, this was not seen with E-ID1. This increase in the enzyme’s concentration would increase the assay’s cost as well; nonetheless, the developed assays are expected to be more accurate in eliminating false-positive results.

A study by (Jamwal et al., 2021) designed the same primers’ sequences, which is expected because the size of targeted *E* gene is small (228 bases), and the same primer designing program was used. However, this is the first study to test this primer set using the colorimetric and fluorometric RT-LAMP methods. Also, the performance of this primer set was improved by optimizing the enzyme concentration and adding reaction enhancers like guanidine hydrochloride. Indeed, after optimization, the detection time of the colorimetric assay was over ten minutes earlier than pre-optimization when testing the same positive samples. It also detected the clinical samples (i.e., J13, J31, J41, and J44) that were previously undetected before increasing the enzyme’s concentration, confirming its effectiveness (Table S1). We tested the effect of Bst 3.0 DNA polymerase compared to the widely used Bst 2.0 DNA polymerase to choose the enzyme that better matches the primers and brings about their best performance. Bst 3.0 DNA polymerase enzyme enhances amplification performance and has a high reverse transcriptase activity, allowing single enzyme RT-LAMP reactions <https://international.neb.com/products/m0374-bst-3-0-dna-polymerase#Product%20Information>. However, the results in Figure S1 indicate that Bst 2.0 enzyme was faster than Bst 3.0 in detecting positive sample triplicates, concluding that this assay has a faster and more efficient detection when Bst 2.0 enzyme is used. Furthermore, the addition of guanidine hydrochloride (GuHCl) to the RT-LAMP reaction has been reported to enhance the speed and sensitivity of SARS-CoV-2 detection. For instance, Zhang et al. reported up to a ten-fold increase in sensitivity and speed in low viral RNA samples; this addition did not increase misamplification in NTC. It is hypothesized that GuHCl enhances base pairing between primers and their targets, significantly improving detection speed and sensitivity (Zhang, 2020b). Another study by Dudley et al. revealed that this addition had lowered the LoD in their assay (Dudley et al., 2020). The improvement was evident with final concentrations between 40-60 mM in a 25 µL reaction volume, with 40 mM being the optimum recommended concentration <https://international.neb.com>\(Zhang et al., 2020b). Most studies reported this enhancement from GuHCl on RNA-extracted clinical samples but not in direct ones (Dudley et al., 2020, Lu et al., 2022). Based on that, we used 40 mM GuHCl in the colorimetric and fluorometric RT-

LAMP reactions; both resulting in 3–7-minute earlier amplifications in positive reactions with GuHCl. Adding GuHCl enhanced the detection speed in this reaction, but it did not improve the sensitivity in detecting the inconclusive sample J43 (positive in RT-qPCR and fluorometric RT-LAMP but negative in colorimetric RT-LAMP). In general, the effect of GuHCl varies depending on the primers used in the reaction. After optimizing the reaction conditions, our assay detects down to 20 copies/µL, or 500 copies/reaction. A study designed an *E* gene primer set that is composed of four primers and had a detection limit of 2000 copies/reaction, which signifies the role of loop primer addition in improving sensitivity. Our LoD is comparable to the one obtained by Zhang et al. (2020a), who designed primer sets that detected down to 480 copies/reaction. Also, another study detected lower copy numbers (10 copies/µL) using primers that target the *ORF1ab* gene (Yu et al., 2020). In general, the LoD depends on the assays’ reaction conditions and the primers’ binding sites.

Generally, the reaction conditions of our developed colorimetric RT-LAMP assay agree with the previous studies that used the same reagents from WarmStart® (New England BioLabs), being 30 minutes incubation time at 65 °C (Amaral et al., 2021, Aoki et al., 2021, Chow et al., 2020, Dao Thi, 2020, de Oliveira Coelho et al., 2021, Huang et al., 2020, Lalli et al., 2021, Luo et al., 2022, Nawattanapaiboon et al., 2021, Promlek et al., 2022). In the fluorometric RT-LAMP assay, we used LavaLAMP™ RNA enzyme (Lucigen), which has an activation temperature of 68-74 °C. Accordingly, based on primers’ melting temperature (T_m_) and after optimizing reaction conditions, 40-60 minutes at 70 °C were the optimum settings.

Both colorimetric and fluorometric assays showed excellent performance in diagnosing COVID-19, being also highly selective against SARS-CoV-2 variants since the primers showed no cross-reactivity against other respiratory viruses *in-silico* and *in-vitro*. The primer set E-ID1 was only used in one study to diagnose SARS-CoV-2 using densitometry and agarose gel electrophoresis to detect RT-LAMP amplification products. However, this primer set did not perform well in their study, so they did not test it further (Jamwal et al., 2021). In addition, their study used Bst 3.0 enzyme for showing the fastest amplification among the other tested enzymes, while our primers performed best when Bst 2.0 enzyme was used. Hence, our optimization to enhance E-ID1 performance in the colorimetric and fluorometric RT-LAMP assay is novel. The assays can be improved by testing alternative enzymes with high strand displacement activity or developing homemade reagents to reduce cost. We found that this assay successfully detected SARS-CoV-2 within 20 minutes. The reaction can be extended up to 120 minutes –without misamplification– in particular for patients with low viral loads. In that regard, the current RT-LAMP test recommends the use of five primers that are rapid and sensitive in detecting COVID-19 infection, especially in symptomatic individuals.

## Conclusion

The purpose of this study is to improve one of the limiting factors of the RT-LAMP technique, which is the non-specific amplifications leading to false-positive results. In general, false-positive results begin to occur within 30 minutes, even after critical primer design. Therefore, current protocols do not recommend exceeding the reaction time over 30 minutes, leading to difficulties in detecting particularly low viral load samples. In order to eliminate the misamplifications, we designed a primer set, namely E-ID1, consisting of five primers that exclude the loop forward (LF) primer. Results showed that the five-primer design provides stable primer targeting and avoids misamplification for 120 minutes. Optimizing the colorimetric and fluorometric RT-LAMP assays decreased the detection time to 20-30 minutes. The colorimetric technique uses only a heating block, without any bulky or expensive instruments, and the test results can be visually interpreted through a simple color change from red to yellow in SARS-CoV-2-infected specimens. Besides, the fluorometric RT-LAMP assay shows a sigmoid (S-shaped) amplification curve in COVID-19-infected samples within 30 minutes. The primers used in this study were originally designed to target a conserved region in the SARS-CoV-2 *E* gene regardless of the variant, and the assays were optimized according to the primer set’s performance. The colorimetric assay yielded 89.5% sensitivity and 97.2% specificity, while sensitivity of 100% and specificity of 96.9 % were obtained in the fluorometric assay when tested on 150 clinical samples. Both assays performed remarkably, making them successful candidates for replacing the conventional RT-qPCR, thereby greatly contributing to improving the economy. The future work would include improving the accuracy and cost of the assays by further optimization or using homemade reagents. Also, the colorimetric RT-LAMP assay has the potential to be developed for in-home use for self-diagnosis. This technique reduces time and abundance on healthcare workers, making it suitable for POC testing in local screening centers, emergency departments, and airports.

## Data Availability

All data produced in the present study are available upon reasonable request to the authors

## Acknowledgements

This study is funded by Institute for Research and Medical Consultations (IRMC) under the project number 2020-IRMC-S-3 and the Deanship of Scientific Research (DSR) of Imam Abdulrahman Bin Faisal University (IAU) Fast track fund of COVID-19 (COVID19-2020-026-IRMC). Huseyin Tombuloglu received the awards.

