## Supplementary Files for "Development of Loop-mediated Isothermal Amplification (LAMP) Assays Using Five Primers Reduces the False-positive Rate in COVID-19 Diagnosis"

**Table S1** Summary of the detection time difference between the fluorometric RT-LAMP assays before and after optimization on some strong and weak positive clinical samples.

| **Samples** | **RT-qPCR (Ct)** | **WarmStart^®^**  **before optimization (minutes)** | **WarmStart^®^**  **after optimization (minutes)** | **LavaLAMP™**  **before optimization (minutes)** | **LavaLAMP™**  **after optimization (minutes)** |
| --- | --- | --- | --- | --- | --- |
| PC mix |  | - | 2  (42 with 2x Bst 2.0) | 37 | 34 |
| Strong positives | | | | | |
| P1 | - | 41 | 30 (2x Bst 2.0) | 35 | 26 |
| P13 | - | 42 | 25 (2x Bst 2.0) | 33 | 27 |
| P14 | - | 37 | 28 (2x Bst 2.0) | 33 | 27 |
| J13 | 24 | Negative | 42 (2x Bst 2.0) | - | 41 |
| J31 | 24 | Negative | 34 | 44 | 37 |
| Weak positives | | | | | |
| J36 | 26 | Negative | Negative | Negative | - |
| J37 | 29 | Negative | Negative | Negative | - |
| J41 | 27 | Negative | 59 | - | 51 |
| J44 | 23 | Negative | 58 | - | 43 |

Abbreviations: RT-qPCR, reverse-transcription quantitative polymerase chain reaction; PC, positive control.

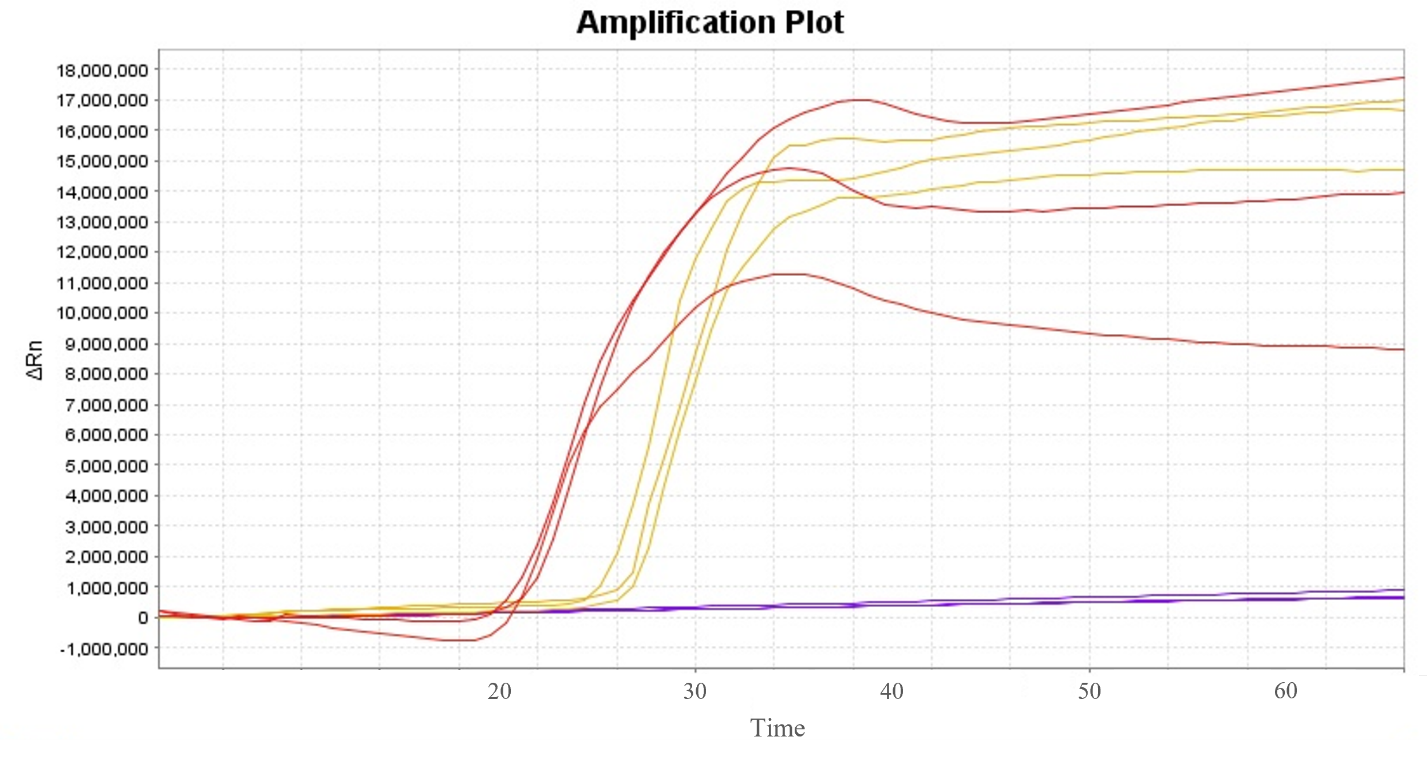

**Figure S1** Comparison between Bst 2.0 (red) and Bst 3.0 (yellow) DNA polymerase performance on three replicates of positive and negative controls (PC and NC) in the fluorometric RT-LAMP using WarmStart^®^ reagents. No improvement was evident in reactions using Bst 3.0 enzyme; thus, Bst 2.0 DNA polymerase has been used for further testing on E-ID1 primers.

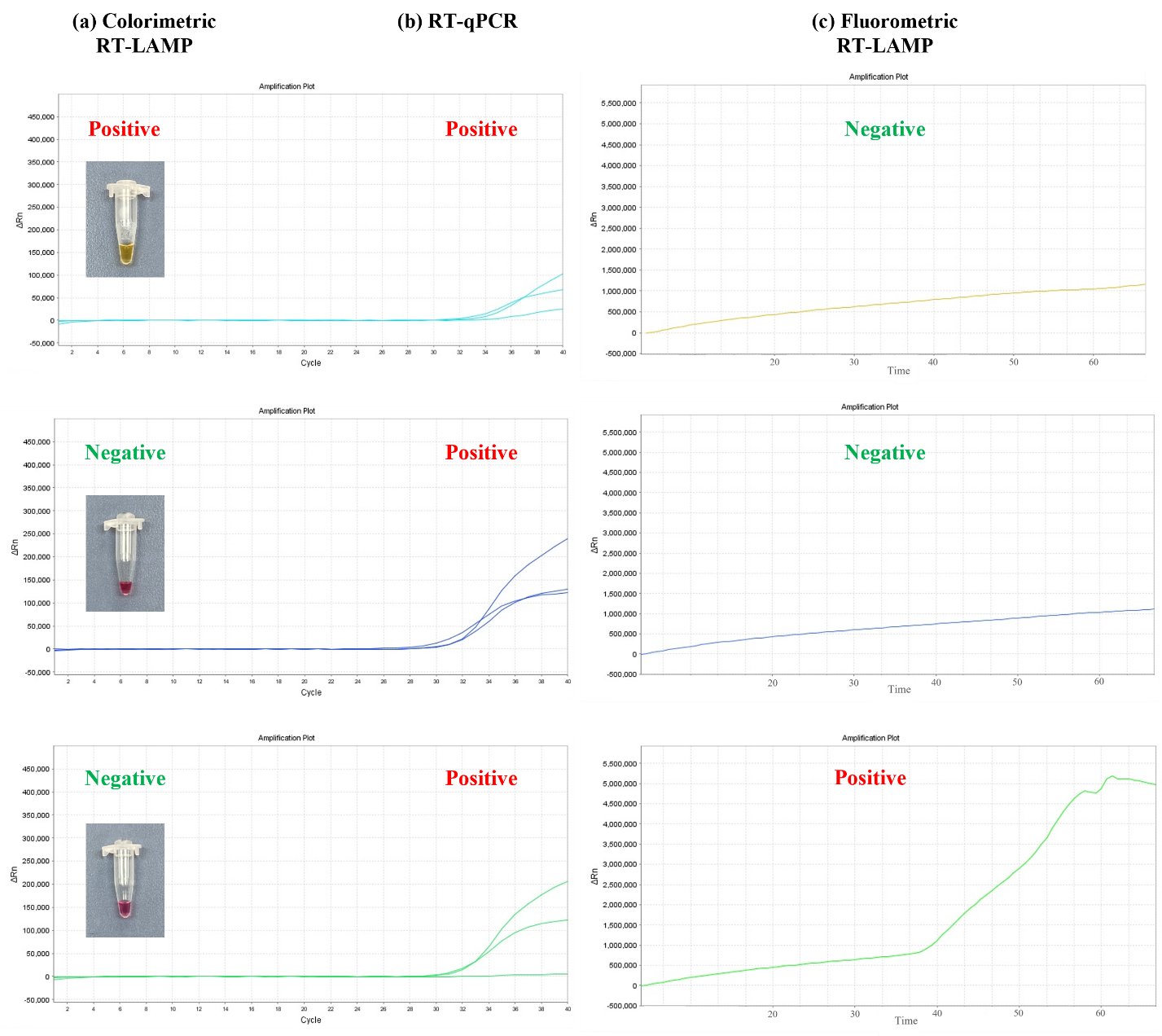

**Figure S2** Comparison of **(a)** colorimetric RT-LAMP, **(b)** RT-qPCR, and **(c)** fluorometric RT-LAMP results of three inconclusive clinical specimens.

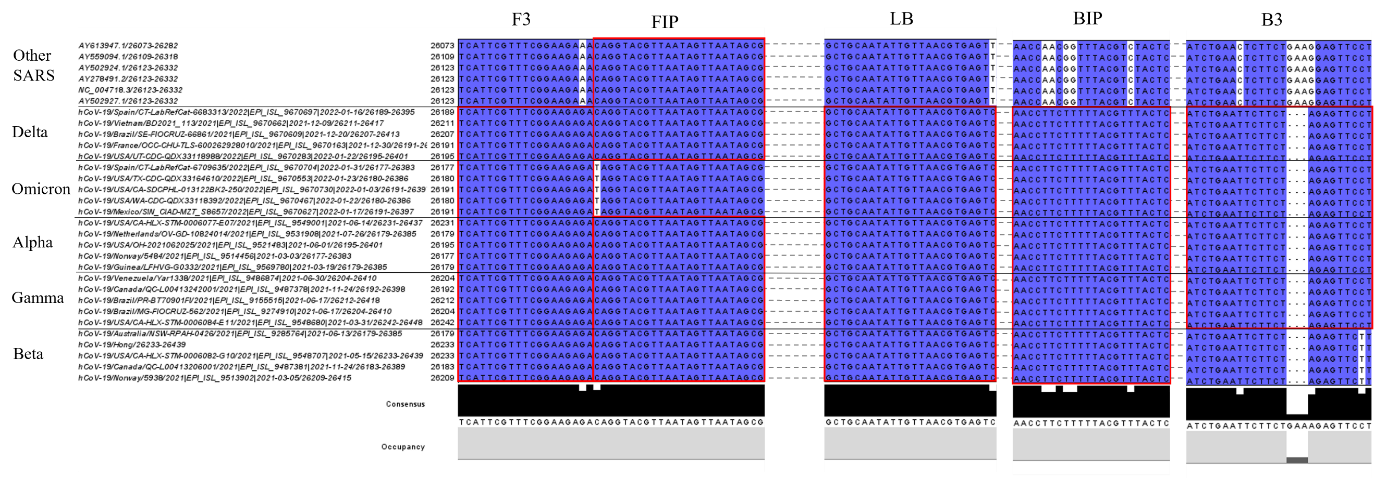

**Figure S3** E-ID1 primers binding sites on the *E* gene of SARS-CoV-2 variants of concern (VOC) and other SARS viruses. This primer sequences bind to a conserved region in SARS-CoV-2 but uncommon for other SARS. Only one point mutation exists in forward inner primer (FIP) binding site.

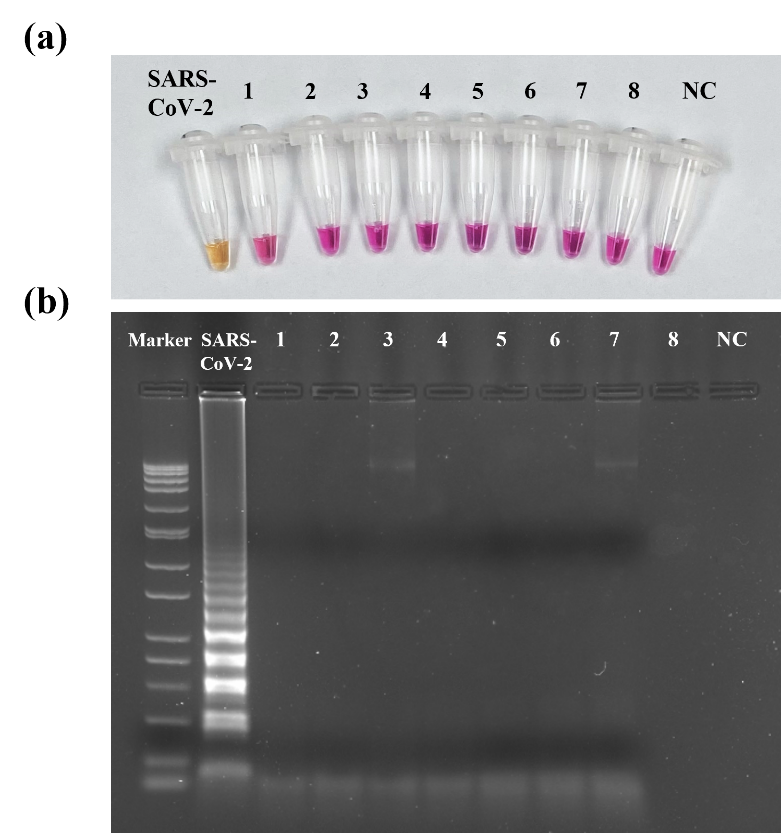

**Figure S4** **(a)** Selectivity of E gene primers colorimetrically tested against clinical samples infected with SARS-CoV-2 along with other respiratory viruses (1-8), these include coronavirus 229 E, parainfluenza virus 3, parainfluenza virus 4, human metapneumovirus A+B, bocavirus, enterovirus, and rhinovirus. The positive color change is seen in SARS-CoV-2 sample only. **(b)** Loading the samples in 2% agarose gel post-reaction showed a ladder-type pattern in COVID-19-infected sample only and nothing in samples with other respiratory viruses, indicating high specificity of the primers against SARS-CoV-2.
